# Dynamics and Development of the COVID-19 Epidemics in the US – a Compartmental Model with Deep Learning Enhancement

**DOI:** 10.1101/2020.05.31.20118414

**Authors:** Qi Deng

## Abstract

**Background:** Compartmental models dominate epidemic modeling. Estimations of transmission parameters between compartments are typically done through stochastic parameterization processes that depend upon detailed statistics on transmission characteristics, which are economically and resource-wide expensive to collect. We apply deep learning techniques as a lower data dependency alternative to estimate transmission parameters of a customized compartmental model, for the purposes of simulating the dynamics of the US COVID-19 epidemics and projecting its further development.

**Methods:** We construct a compartmental model. We develop a multistep deep learning methodology to estimate the model’s transmission parameters. We then feed the estimated transmission parameters to the model to predict the development of the US COVID-19 epidemics for 35 and 42 days. Epidemics are considered suppressed when the basic reproduction number (*R*_0_) becomes less than one.

**Results:** The deep learning-enhanced compartmental model predicts that *R*_0_ will become less than one around June 19 to July 3, 2020, at which point the epidemics will effectively start to die out, and that the US “Infected” population will peak round June 18 to July 2, 2020 between 1·34 million and 1·41 million individual cases. The models also predict that the number of accumulative confirmed cases will cross the 2 million mark around June 10 to 11, 2020.

**Conclusions:** Current compartmental models require stochastic parameterization to estimate the transmission parameters. These models’ effectiveness depends upon detailed statistics on transmission characteristics. As an alternative, deep learning techniques are effective in estimating these stochastic parameters with greatly reduced dependency on data particularity.

## Introduction

The COVID-19 pathogen that ravages China, Europe and the US since December 2019 is a member of the coronavirus family, which also includes the SARS-CoV and MERS-CoV. In the US, as of June 4, 2020, there have been 1,872,660 confirmed cases and 108,211 deaths.

The COVID-19 pandemic is still in progress, and most of the noticeable early research is of descriptive nature, focusing on reported cases to establish the baseline demographic parameters for the disease, such as age, gender, health and medical conditions, in addition to the disease’s clinical manifestations, in a Chinese context. These studies includes reports on demographic characteristics, epidemiological and clinical characteristics, exposure and travel history to the epicenter, illness timelines of laboratory-confirmed cases^1–5^, as well as epidemiological information on patients from social networks, local, national and international health authorities^6^. The spread of SARS-CoV-2 outside of China (e.g. Iceland) is also analyzed^7^, albeit limitedly. From a US perspective, concerned with the worsening situation in New York City, researchers characterize information on the first 393 consecutive Covid-19 patients admitted to two hospitals in that city^8^.

Some stage-specific studies on COVID-19 patients have also been carried out, including a single-centered, retrospective study on critically ill adult patients in Wuhan, China^9^, and a retrospective, multi-center study on adult laboratory-confirmed inpatients (≥18 years old) from two Wuhan hospitals who have been discharged or have died^10^.

### COVID-19 epidemic modeling

There have been attempts to model the COVID-19 epidemic dynamics. These studies all add a worldwide mobile dimension, reflecting a higher level of mobility and globalization in 2020 than 2003 (SARS) and even 2013 (MERS). The SEIR model is used to infer the basic reproduction ratio, and simulate the Wuhan epidemic^11^; it considers domestic and international air travel to/from Wuhan to other cities to forecast the national and global spread of the virus. More sophisticated models have also been developed to correlate risk levels of foreign countries with their travel exposure to China^12–13^, including a stochastic dual-SEIR approach on both Wuhan population and international travelers to estimate how transmission have varied over time from Wuhan to international destinations^13^. Simulations on the international spread of the COVID-19 after the start of travel ban from Wuhan on Jan 23, 2020 have also been conducted^14^, which apply the Global Epidemic and Mobility Model (GLEAM) to a multitude of Chinese and international cities, and a SEIR variety (SLIR) to project the impact of human-to-human transmissions. To simulate the transmission mechanism itself, a Bats-Hosts-Reservoir-People (BHRR) network is developed to simulate potential transmission from the infection sources (i.e., bats) to humans^15^.

Since March 2020, with the COVID-19 outbreak winding down in China, researchers have dedicated more efforts in analyzing the effectiveness of containment measures. Mobility and travel history data from Wuhan is used to ascertain the impact of the drastic control measures implemented in China^16^. A research investigates the spread and control of COVID-19 among Chinese cities with data on human movements and public health interventions^17^. Utilizing the contact data for Wuhan and Shanghai and contact tracing information from Hunan Province, a group of researchers build a transmission model to study the impact of social distancing and school closure18.

### Theoretical Foundation

Compartmental models dominate epidemic modeling on COVID-19 epidemics (and previous coronavirus outbreaks), and they require detailed statistics on transmission characteristics to estimate the stochastic transmission parameters between compartments. Essentially, these models correlate factors such as geographic distances and contact intensities among heterogeneous subpopulations with gradient probability decay. Technically, transmission parameterization applies Bayesian inference methods, such as Marcov Chain Monte Carlo (MCMC) or Gillespie algorithm^19^ simulations to form probability density functions (PDFs) on cross-section, in order to estimate parameters for each timestep of a multivariate time series construct. These detailed statistics on transmission characteristics are economically and resource-wide expensive to collect.

We are particularly interested in extended compartmental models that cover multiple inter-connected and heterogeneous subpopulations^10,15,20^. There are also some pure time series analyses on epidemic dynamics outside of the compartmental modeling mainstream, for example, the AutoRegressive Integrated Moving Average (ARIMA) approach^21^ that is typically found in financial applications. They provide another perspective.

We develop a multistep, multivariate deep learning methodology to estimate the transmission parameters. We then feed these estimated transmission parameters to a customized compartmental model to predict the development of the US COVID-19 epidemic.

We establish a SEIR-variety discrete time series on a daily interval as the theoretical foundation for a deep learning-enhanced compartment model. We start with the construction of a so called SEIRQJD (SEIR-Quarantined-Isolated-Deceased) model (Fig. 1).

**Fig. 1.**
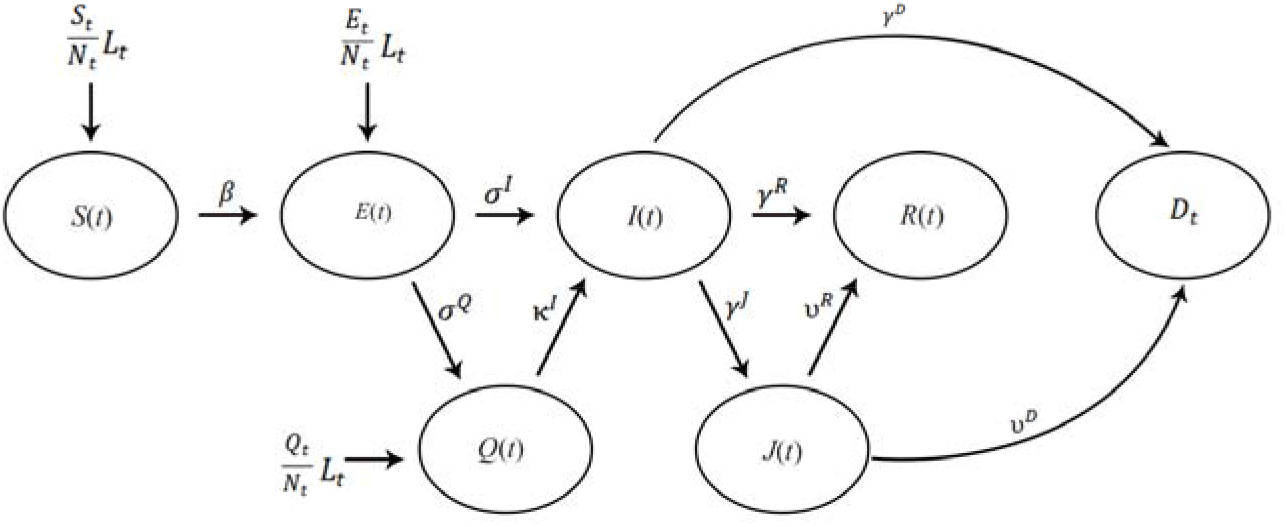
The SEIRQJD Model

We use the US COVID-19 epidemic datasets from John Hopkins University Center for Systems Science and Engineering (JHU CSSE) Github COVID-19 data depository, which do not include direct Exposed (E) and quarantined (Q) data, we set all transmission parameters to/from the “E” and “Q” compartment (,,) to zero. Furthermore, the datasets assume that all Deaths (D) arise from the Isolated population (J), thus we also set the transmission parameter from Infectious (I) to Deceased (D),, to zero. We then simplify the SEIRJD model to a SIRJD (SIR-Isolated-Deceased) construct, in which a population is grouped into five compartments:

1. Susceptible (S): The susceptible population arises at a percentage (−) of a net influx of individuals ().
2. Infectious (I): The infectious individuals are symptomatic, come from the Susceptible compartment, and further progress into the Isolated or Recovered compartments.
3. Isolated (J): The isolated individuals have developed clinical symptoms and have been isolated by hospitalization or other means of separation. They come from the Infectious compartment and progress into the Recovered or Deceased compartments
4. Recovered (R): The recovered individuals come from Infectious and Isolated compartments and acquire lasting immunity (there has yet any contradiction against this assumption).
5. Deceased (D): The deceased cases come from Infectious and Isolated compartments.

The SIRJD model has a daily (∆*t* = 1) multivariate time series construct given by the follow matrix form:

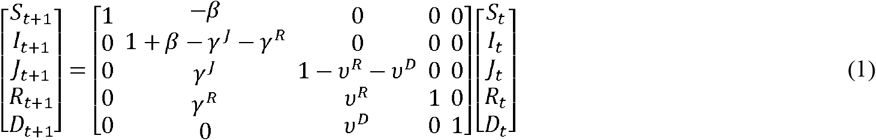

or:

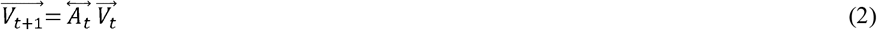

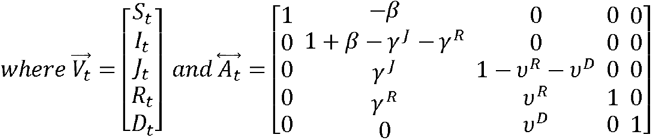

The Greek letters in the time series are transmission parameters defined in the state diagram in Figure 1. Essentially, all these parameters are stochastic.

Since we need to estimate the transmission parameters, we can rewrite and rearrange Equations (1) and (2) to the following matrix representation:

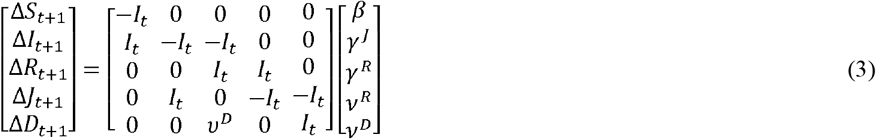

or:

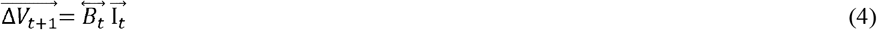

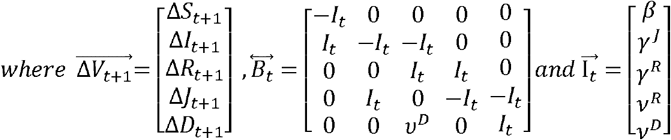

### Data

We collect the following US COVID-19 datasets from the JHU CSSE data depository^➀^.

1. Dataset 1: The JHU CSSE updates daily records (confirmed, active, dead, recovered, hospitalized, etc.) from April 12, 2020. We use these detailed case data to construct the compartmental model.
2. Dataset 2: The JHU CSSE updates two time series on daily basis. One tracks confirmed cases and the other dead cases, both starting January 22, 2020. We use the confirmed/dead cases to form training data for deep learning.

The JHU CSSE dataset has an almost precise period of 7 days (±1 days), reflecting that a majority of the reporting agencies in the country choose to update their respective statistics on a weekly, fixed-calendar interval. We run a 7-day moving average on the dataset to smooth out this “unnatural” data seasonality.

### Methodology

We then conduct the following step-by-step operations, to model the US epidemic:

1. We construct an in-sample SIRJD time series starting April 12, 2020 with Dataset 1.
2. We use the in-sample SIRJD time series constructed in Step 1 to come up with an in-sample time series for the two most critical daily transmission parameters (*β* and *γ^R^*).
3. We construct a confirmed/dead-case time series starting from January 22, 2020 (in-sample) with Dataset 2.
4. We apply two deep learning approaches, the standard DNN (Deep Neural Networks) and the advanced RNN-LSTM (Recursive Neural Networks – Long Short Term Memory), to fit the confirmed/dead in-sample time series from Step 3, and predict the further development of confirmed/dead cases for 35 and 42 days (out-of-sample).
5. We use the confirmed/dead in-sample time series from Step 3 as training data and the in-sample *β* and *γ^R^* time series from Step 2 as training label, and apply the DNN and RNN-LSTM techniques to predict *β* and *γ^R^* for 35 and 42 days (out-of-sample).
6. Finally, we use the predicted (out-of-sample) transmission parameters (*β* and *γ^R^*) from Step 5 to simulate 35-day and 42-day progressions (out-of-sample) of the SIRJD model (particularly the SIR portion) in a recursive manner, starting with the data point of the last timestep from the in-sample SIRJD time series from Step 1.

Fig. 2 is the flowchart to illustrate the dataset and methodology.

**Fig. 2.**
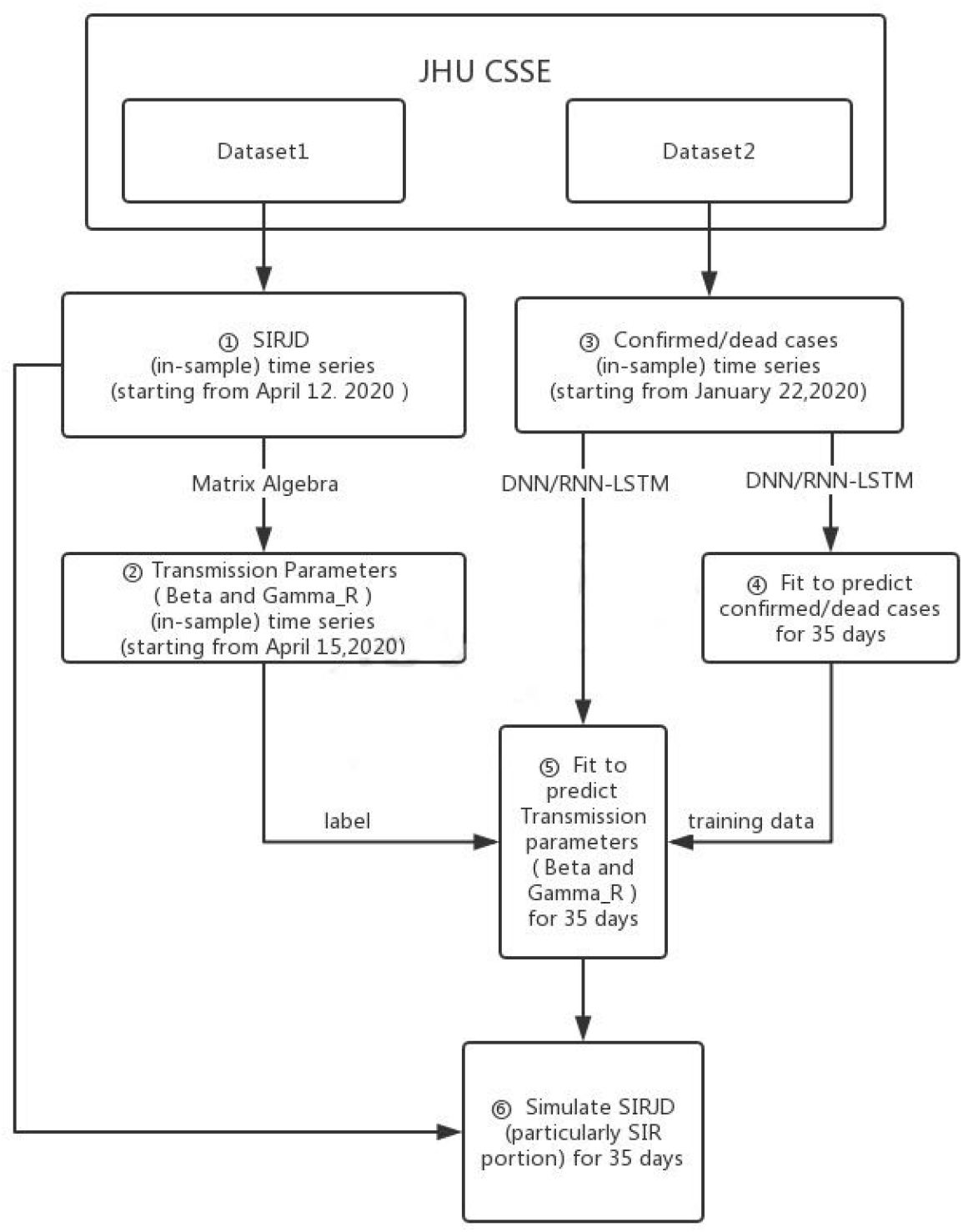
The dataset and methodology

### Results

The results are illustrated in Figs. 3–6 (35-day forecast) and Figs. 7–10 (42-day forecast).

**Fig. 3.**
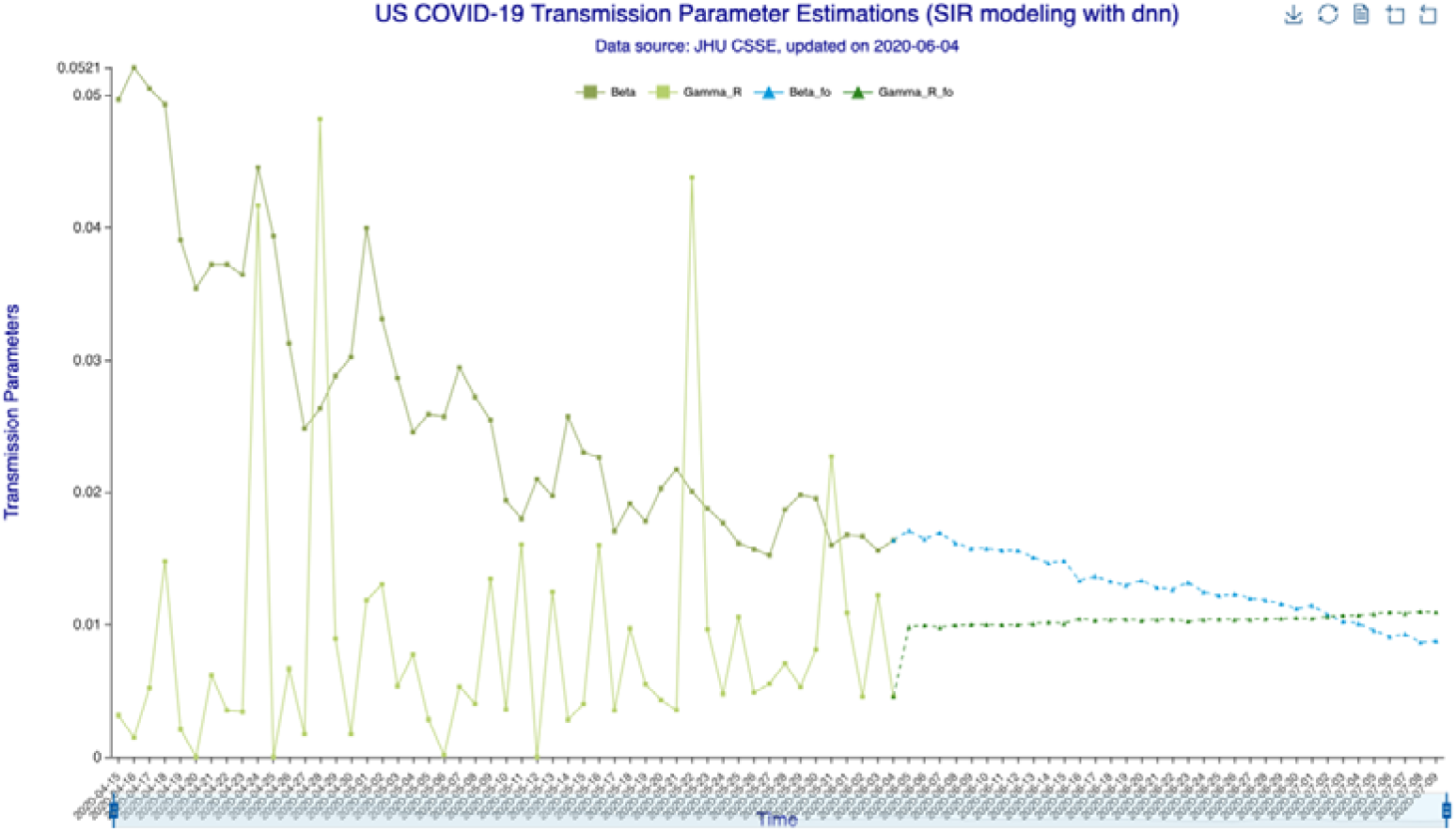
The results – transmission parameter estimations (DNN) for 35 days Beta is the “Susceptible-to-Infected” transmission parameter () and Gamma_R is the “Infected-to-Recovered” transmission parameter () for the in-sample (observed) data. Beta_fo is the forecasted and Gamma_R_fo is forecasted for the out-of-sample (forecasted) data.

**Fig. 4.**
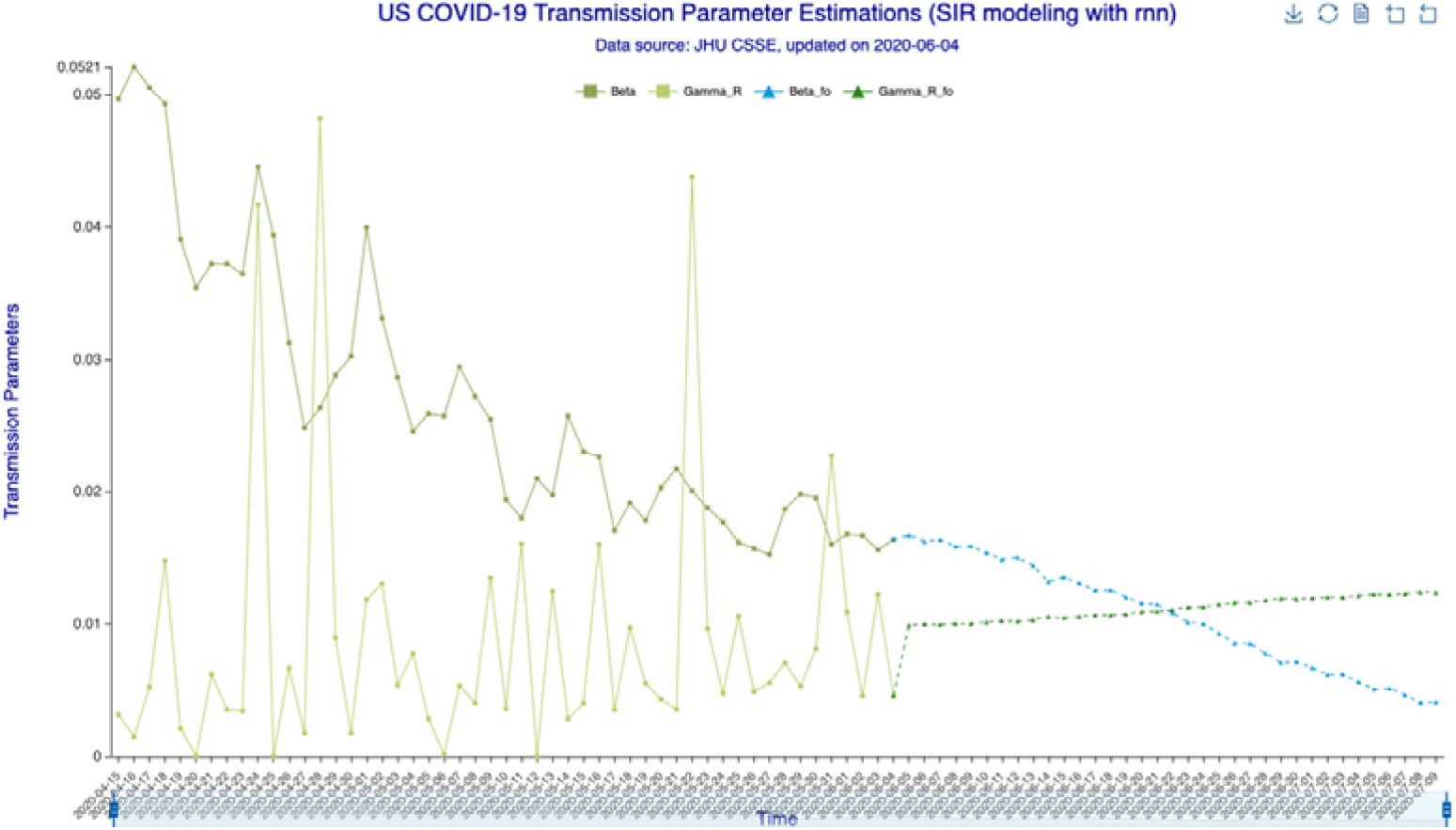
The results – transmission parameter estimations (RNN-LSTM) for 35 days Beta is the “Susceptible-to-Infected” transmission parameter () and Gamma_R is the “Infected-to-Recovered” transmission parameter () for the in-sample (observed) data. Beta_fo is the forecasted and Gamma_R_fo is forecasted for the out-of-sample (forecasted) data.

**Fig. 5.**
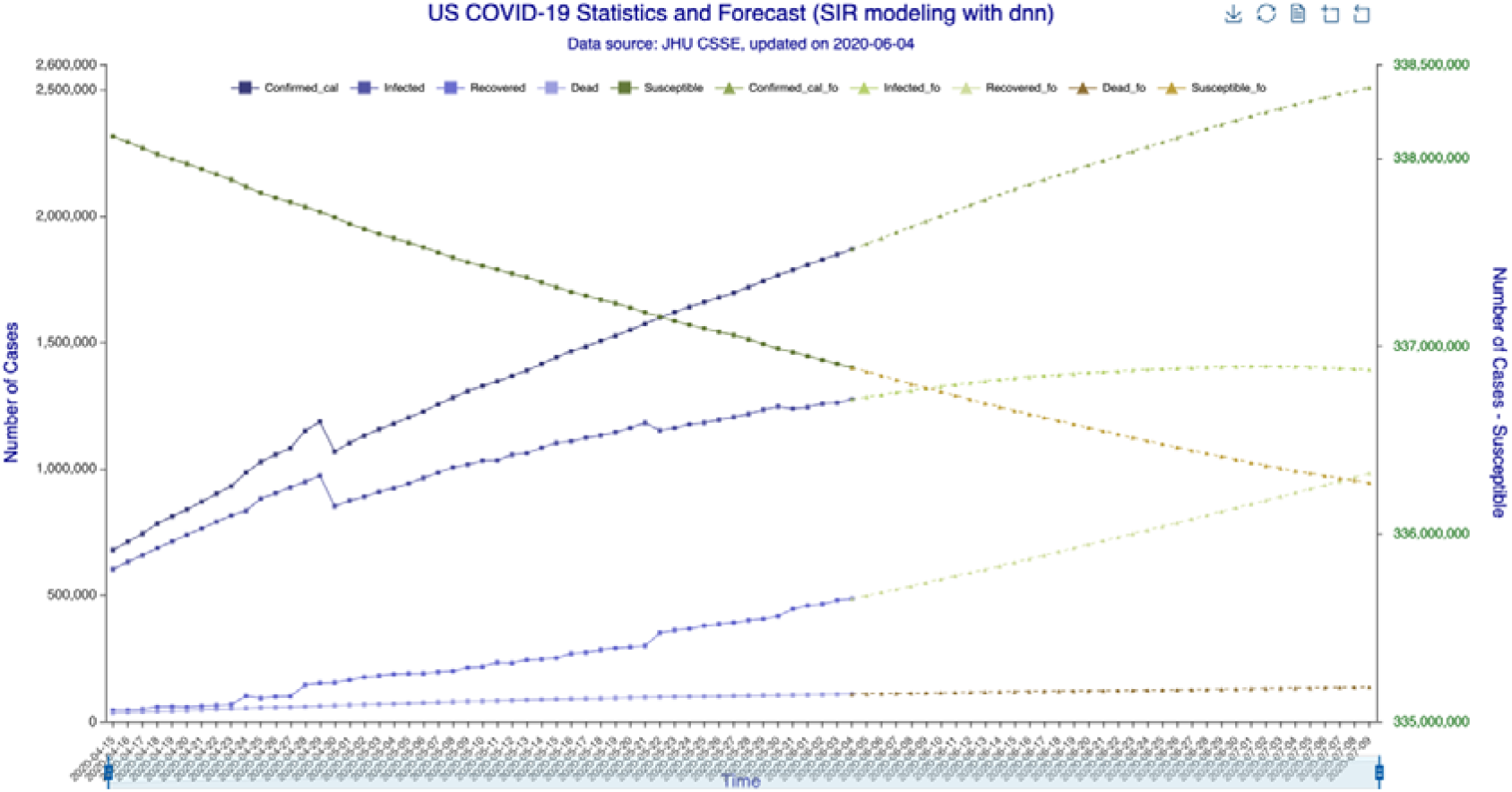
The results – SIR model forecasting (DNN) for 35 days Susceptible, Infected, Recovered, Dead are in-sample compartmental model data, Confirmed_cal is the in-sample number of confirmed cases. Susceptible_fo, Infected_fo, Recovered_fo, Dead_fo and Confirmed_cal_fo are their out-of-sample (forecasted) counterparts. The right y-axis is for Susceptible/Susceptible_fo, while the left y-axis is all others. The right y-axis is needed for scaling purpose, as Susceptible/Susceptible_fo are derived from the total population.

**Fig. 6.**
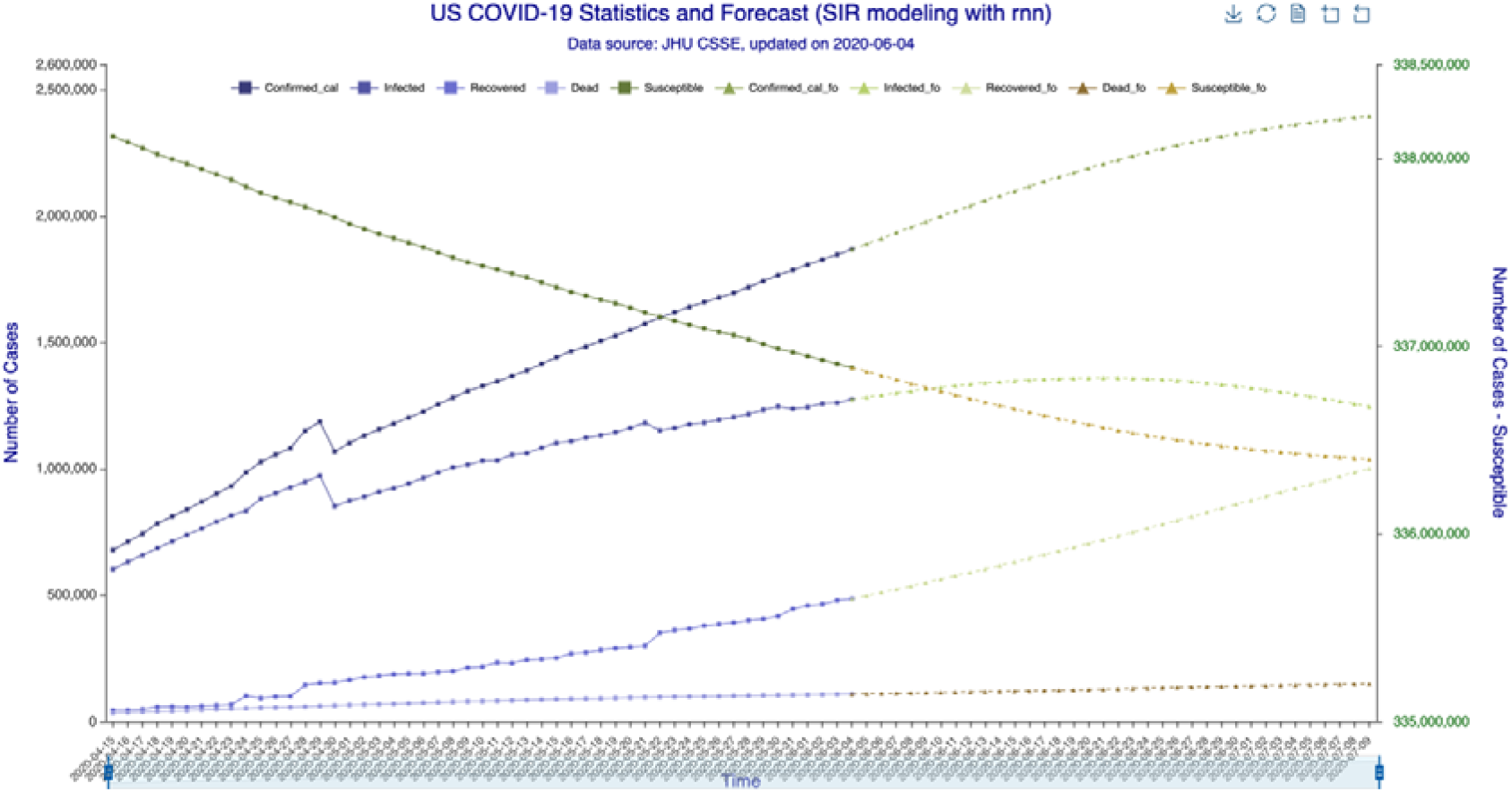
The results – SIR model forecasting (RNN-LSTM) for 35 days Susceptible, Infected, Recovered, Dead are in-sample compartmental model data, Confirmed_cal is the in-sample number of confirmed cases. Susceptible_fo, Infected_fo, Recovered_fo, Dead_fo and Confirmed_cal_fo are their out-of-sample (forecasted) counterparts. The right y-axis is for Susceptible/Susceptible_fo, while the left y-axis is all others. The right y-axis is needed for scaling purpose, as Susceptible/Susceptible_fo are derived from the total population.

**Fig. 7.**
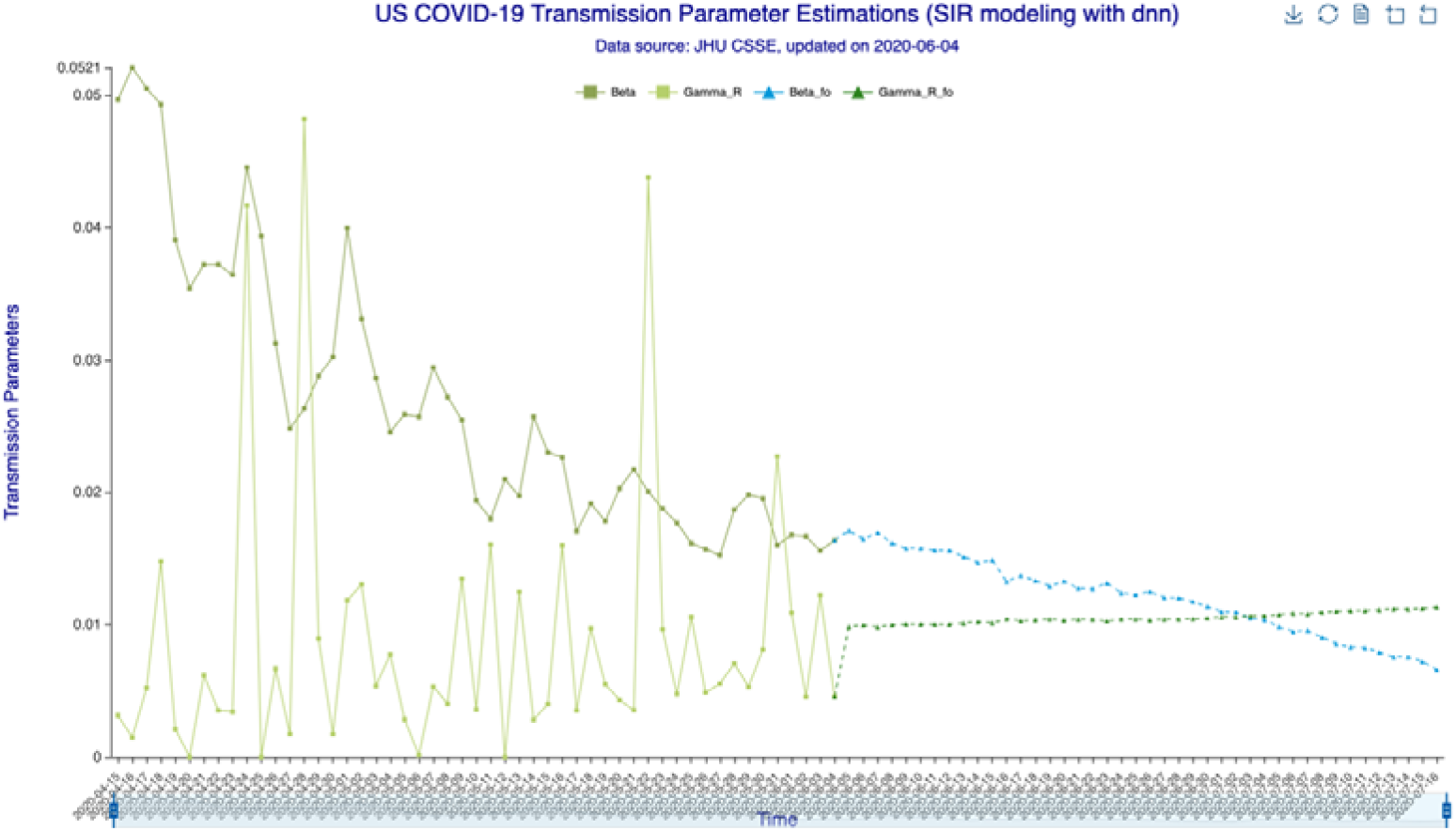
The results – transmission parameter estimations (DNN) for 42 days Legends same as Fig. 3.

**Fig. 8.**
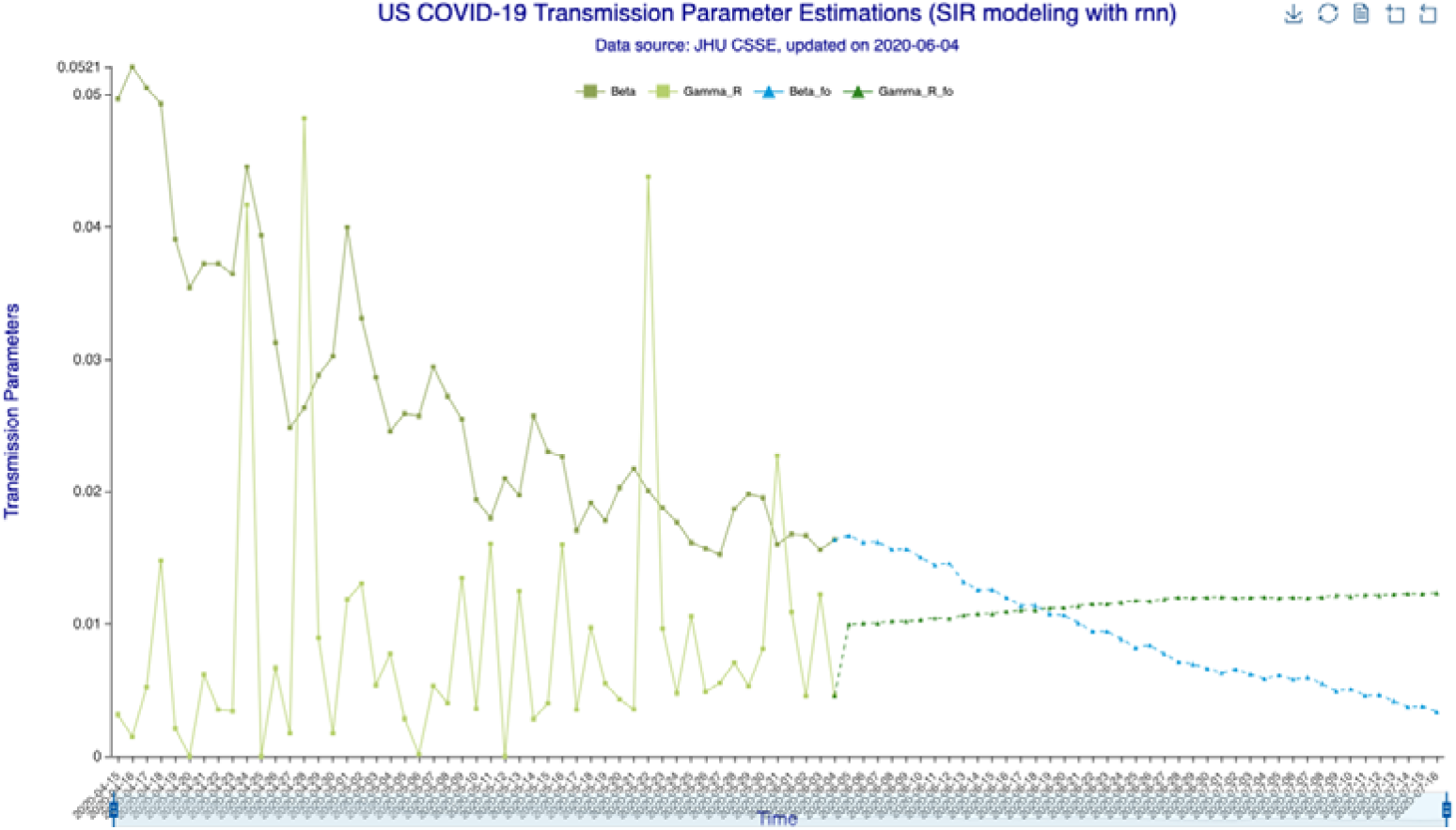
The results – transmission parameter estimations (RNN-LSTM) for 42 days Legends same as Fig. 4.

**Fig. 9.**
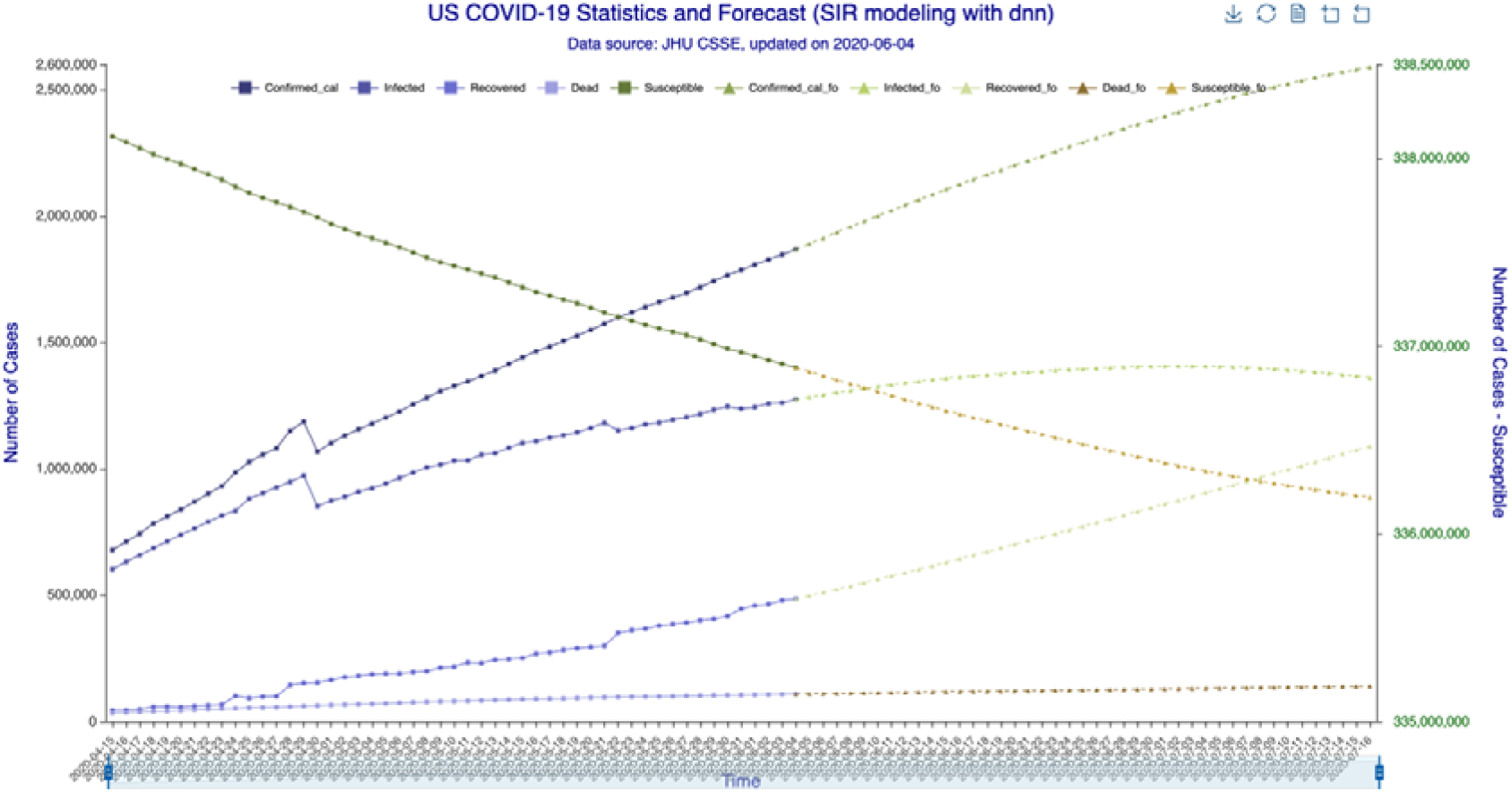
The results – SIR model forecasting (DNN) for 42 days Legends same as Fig. 5.

**Fig. 10.**
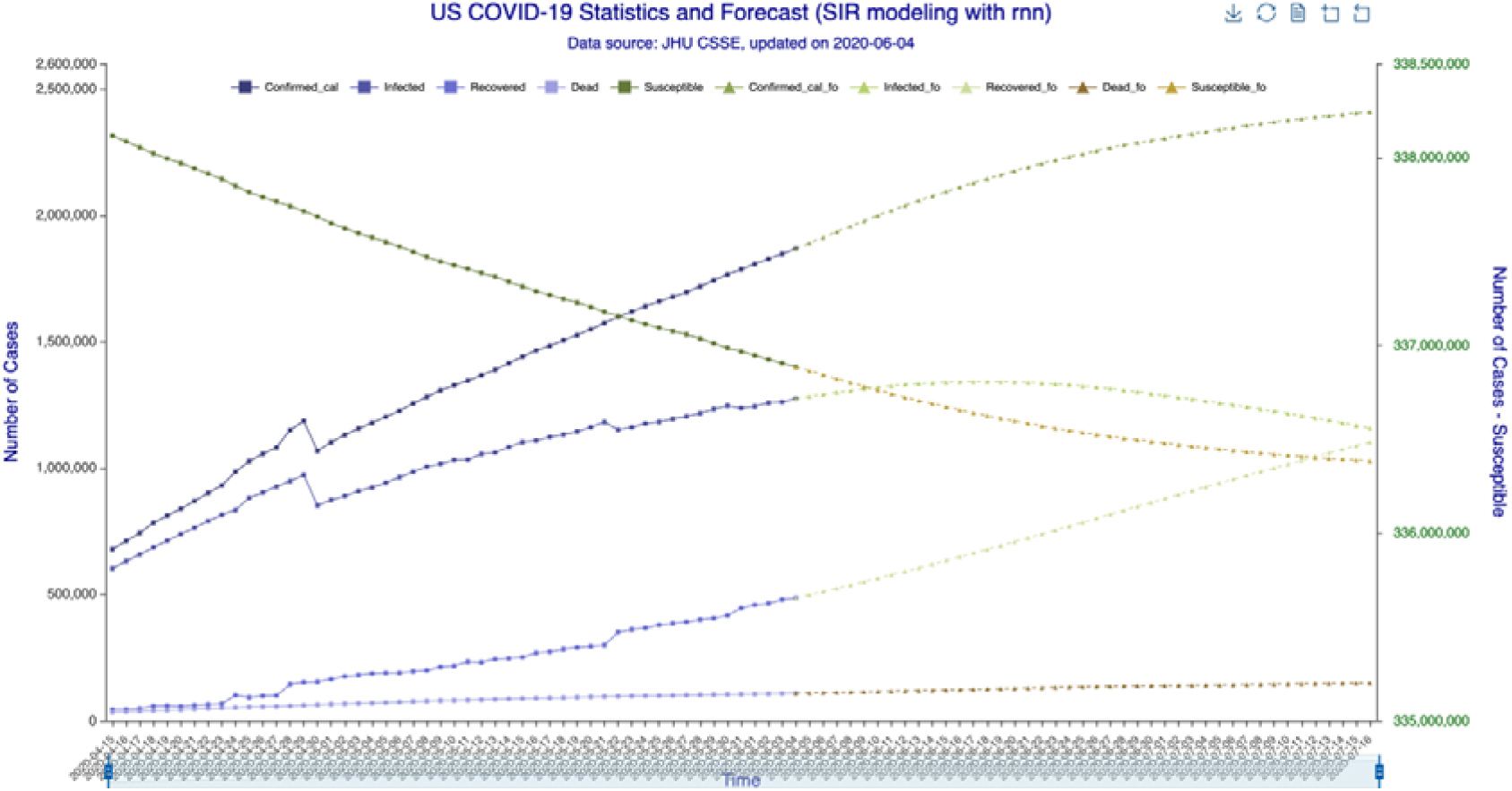
The results – SIR model forecasting (RNN-LSTM) for 42 days Legends same as Fig. 6.

In Fig. 3 (35-day forecast), the DNN method predicts on July 3, 2020, the “Infected-to-Recovered” transmission parameter will rise above and stay above the “Susceptible-to-Infected” transmission parameter. This means that the value of the basic reproduction rate,, will become less than one and that the spread of COVID-19 in the US will effectively end on that day. In Fig. 4 (35-day forecast), the RNN-LSTM method gives a more aggressive prediction that *γ^R^* will overtake *β* on June 22, 2020. Thus, with 35-day forecast, we predict that the tide of the US epidemic will turn around June 22 to July 3, 2020 timeframe.

In Fig. 5 (35-day forecast), the DNN method predicts that the US “Infected” population will peak on July 2, 2020 at 1,406,136 individual cases. In Fig. 6 (35-day forecast), the RNN-LSTM method predicts that the US “Infected” population will peak on June 21, 2020 at 1,359,619. For 35-day forecast, the deep learning methods predict that the number of accumulative confirmed cases will cross the 2 million mark on June 10, 2020 at 2,000,525 by DNN (Fig. 5) and on June 11, 2020 at 2,019,923 by RNN-LSTM (Fig. 6).

In Fig. 7 (42-day forecast), the DNN method gives the same prediction (as 35-day forecast) that *γ^R^* will overtake *β* on July 3, 2020. While in Fig. 8 (42-day forecast), the RNN-LSTM method gives a more aggressive (than 35-day forecast) prediction that *R*_0_ will become less than one on June 19, 2020. Comparing Figs. 7–8 with Figs. 3–4, we observe that the 42-day forecast gives a wider range (June 19 to July 3, 2020) than the 35-day forecast (June 22 to July 3, 2020) on turnaround timeframe. The reason is that, for the same (in-sample) training data size, longer forecast produces wider probability distributions.

In Fig. 9 (42-day forecast), the DNN method predicts that the US “Infected” population will peak on July 2, 2020 at 1,406,404 individual cases. In Fig. 10 (42-day forecast), the RNN-LSTM method predicts that the US “Infected” population will peak on June 18, 2020 at 1,341,021. For 42-day forecast, the deep learning methods predict that the number of accumulative confirmed cases will cross the 2 million mark on June 10, 2020 at 2,000,585 by DNN (Fig. 9) and on June 11, 2020 at 2,018,776 by RNN-LSTM (Fig. 10), which are consistent with the 35-day forecasts.

## Discussion

We apply DNN and RNN-LSTM techniques to estimate the stochastic transmission parameters for a SIRJD model with a discrete time series construct. We then use the predicted transmission parameters to forecast the further development of the US COVID-19 epidemics.

We make use of two US COVID-19 datasets from the JHU CSSE data depository. The first dataset includes detailed daily records (confirmed, active, dead, recovered, hospitalized, etc.) starting from April 12, 2020, from which we construct the SIRJD model. The second dataset includes time series tracking confirmed and dead cases starting from January 22, 2020, which we use to construct training data for deep learning. The JHU CSSE data has an almost precise period of 7 days (±1 days) that masks the true epidemic dynamics, thus we run a 7-day moving average on the dataset to smooth out this data seasonality.

We then apply DNN and RNN-LSTM deep learning techniques to fit the confirmed/dead series to predict the further development of confirmed/dead cases, as well as to predict the “Susceptible-to-Infected” and “Infected-to-Recovered” transmission parameters (*β* and *γ^R^*) for 35 and 42 days. Finally, we use the predicted transmission parameters (*β* and *γ^R^*) from simulate the epidemic progression for 35 and 42 days.

The deep learning implementations for 35-day forecast predict that the basic reproduction rate (*R*_0_) will become less than one around June 22 to July 3, 2020, and for 42-day forecast around June 19 to July 3, 2020, at which point the spread of the coronavirus will effectively start to die out.

The implementations for 35-day forecast predict that the US “Infected” population will peak around June 21 to July 2, 2020, between 1,359,619 and 1,406,136 individual cases, respectively. The implementations for 42-day forecast predict that the peak will arrive on June 18 to July 2, 2020, between 1,341,021 and 1,406,404 individual cases.

All implementations indicate that the number of accumulative confirmed cases will cross the 2 million mark around June 10 to 11, 2020.

With the introduction of the deep learning-enhanced compartmental model, we provide an effective and easy-to-implement alternative to prevailing stochastic parameterization, which estimates transmission parameters through either probability likelihood maximization, or Marcov Chain Monte Carlo simulation. The effectiveness of the prevalent approach depends upon detailed statistics on transmission characteristics among heterogeneous subpopulations, and such statistics are economically and resource-wide expensive. On the other hand, deep learning techniques uncover hidden interconnections among seemly less related data, reducing prediction’s dependency on data particularity. Future research on deep learning’s utilities in epidemic modeling can further enhance its forecasting power.

## Data Availability

Up-to-date time series data available up request.

## Acknowledgements

The authors thank Ms. Liu Chang and Mr. Liu Shuigeng with Cofintelligence Fintech Co, Ltd. (Hong Kong and Shanghai) for data collection and formatting. Authors declare no competing interests.

## Supplementary Materials

Dataset_1 time series (constructed): covid_us.csv

Dataset_2 time series (constructed): covid_us_ts.csv

## Footnotes

➀ The JHU CSSE Github COVID-19 data depository link is at https://github.com/CSSEGISandData/COVID-19.

## Notes

### Competing Interest Statement

The authors have declared no competing interest.

